# Statistical Analyses of the Public Health and Economic Performance of Nordic Countries in Response to the COVID-19 Pandemic

**DOI:** 10.1101/2020.11.23.20236711

**Authors:** Daniel V. Gordon, R. Quentin Grafton, Stein Ivar Steinshamn

**Affiliations:** University of Calgary University of Stavanger; The Australian National University; Department of Business and Management Science/Centre for Applied Research at NHH Norwegian School of Economics (NHH)

**Keywords:** social distancing, difference-in-differences, health-economic trade-offs, Sweden

## Abstract

**Aim:** To compare trends and undertake statistical analyses of differences in public health performance (confirmed cases and fatalities) of Nordic countries; Denmark, Finland, Norway and Sweden, and New Zealand, in response to the COVID-19 pandemic.

**Methods:** Per capita trends in total cases and per capita fatalities were analysed and difference-in-difference statistical tests undertaken to assess whether differences in stringency of mandated social distancing (SD) measures, testing rates and border closures explain cross-country differences.

**Results:** Sweden is a statistical outlier, relative to its Nordic neighbours, for both per capita cases and per capita fatalities associated with COVID-19 but not in terms of the reduction in economic growth. Sweden’s public health differences, compared to its Nordic neigbours, are partially explained by differences in terms of international border closures and the level of stringency of SD measures (including testing) implemented from early March to June 2020.

**Conclusions:** We find that: one, early imposition of full international travel restrictions combined with high levels of government-mandated stringency of SD reduced the per capita cases and per capita fatalities associated with COVID-19 in 2020 in the selected countries and, two, in Nordic countries, less stringent government-mandated SD is *not* associated with higher quarterly economic growth.

## Introduction

The SARS-CoV-2 virus [1] that causes COVID-19 was first observed in China in December 2019 and declared a global pandemic on 11 March 2020 [2]. As early as February 2020, some countries implemented a series of public health measures to stop the spread of COVID-19. Measures differed by country but included international travel restrictions from China, and later from other infected\ countries, as well as quarantine of travellers from COVID-19 ‘hot spots’, and the testing for the virus in new arrivals who exhibited fever or flu-like symptoms.

The European Union (EU) first imposed internal border restrictions on persons travelling from Italy on 17 February 2020. Several EU member countries also imposed their own border controls. On 17 March, the EU imposed external border closures for all non-essential travel. By April 2020, most high-income countries had imposed some form of border restrictions while some countries (such as Australia and New Zealand) further required that all incoming arrivals be placed in supervised quarantine for 14 days and to test negative for virus shedding before leaving quarantine.

Unlike its Nordic neighbours, Sweden maintained an open border within the EU and beyond and imposed no quarantine requirements for arrivals. From 31 October 2020, Sweden banned non-essential travel, except for Swedish citizens, from countries outside of the EU [3]. Sweden’s open border policy arose because, according to Anders Tegnell in April 2020, and who is the Chief Epidemiologist at the Swedish Public Health Agency, “Closing borders, in my opinion, is ridiculous, because COVID-19 is in every European country now. We have more concerns about movements inside Sweden.” [4].

In addition to an open border, Sweden has minimised its government-mandated social distancing (SD) measures to suppress COVID-19 infections [5] and relied, instead, primarily on voluntary behaviours of Swedish residents to comply with national health advisories [6]. At least in the early months of the pandemic, voluntary SD reduced the frequency and proximity of social interactions outside of the home by Swedes [7]. Nevertheless, both the number of confirmed COVID-19 cases and fatalities in Sweden were substantially higher in the period March to June 2020 than in its Nordic neighbours that adopted government-mandated SD to suppress infections.

The justification for the reliance on voluntary SD and advisories in Sweden, rather than enforcement of government mandated SD rules, was two-fold. First, it was claimed that government-mandated and stringent SD would not be successful over the long term, should the pandemic continue for years, because its population would not accept or comply with extended periods of mandated SD [6]. Second, voluntary SD to control COVID-19 infections would impose a lower cost on the economy than stringent government-mandated SD [8].

Here, we evaluated the public health and economic costs of the Sweden’s virtual lockdown to government-mandated lockdowns with its Nordic neighbours, and New Zealand. Our analysis used public data on COVID-19 in terms of; confirmed cases, fatalities, testing, the stringency of mandated SD measures (including border closures), to determine *if* there is a statistical difference between the approach taken by Sweden versus that of its Nordic neighbours (Denmark, Finland and Norway), and New Zealand, in relation to public health and economy growth. Our approach was to evaluate trends over time in per capita cases, per capita fatalities, per capita testing, economic costs (as measured by change in Gross Domestic Product, GDP), and stringency of mandated SD measures in all countries and, where possible, to test for statistical differences among the countries.

## Methods

### Data sources

Publicly available national and global data sources from 1 January to 31 October 2020 were used to provide time trends of cases, fatalities, testing, quarterly GDP growth and stringency of government-mandated SD measures [9] for the Nordic countries; Denmark, Finland, Norway and Sweden, and also New Zealand. All data sources and summary comparative statistics are available on request from the authors.

### Study design

The four Nordic countries are peers because of similarities in terms of age structure, life expectancy, welfare support, geography, population density, per capita income, and level (hospital beds per capita) and quality of health care. New Zealand was also included as a comparator because, unlike Sweden, it adopted a ‘go hard and go early’ strategy that included highly restrictive border controls, 14-days quarantine of all arrivals and other stringent government-mandated SD measures.

Unlike many other countries, the explicit aim of the New Zealand government was to eliminate community transmission [10] and which it achieved twice; first, in an initial and larger outbreak from March to May [11] and again in a much smaller outbreak from August-October 2020. The success of New Zealand’s elimination strategy is such that it actually *reduced* its mortality rate over the period January to May 2020, a period that coincided with its first outbreak, relative to previous years [12].

### Statistical analyses

Given there are only five countries in terms of our comparisons, standard statistical tests of differences in means with respect to confirmed cases, fatalities, testing, stringency of mandated SD measures, and changes in GDP are not valid. Instead, time-trend comparisons are provided on the selected variables for the four Nordic countries and New Zealand from 1 January to 31 October 2020.

A difference in difference (DiD) analysis was separately undertaken using daily data controlling for monthly fixed effects for the period March to October 2020 in relation to two dependent variables; one, per capita confirmed cases and, two, per capita COVID-19 fatalities for the four Nordic countries. The DiD method is widely used to test whether or not there is a statistically significant difference in data [13, 14]. Dummy variables were included for each country (Norway is the reference country) to account for unaccounted cross-country differences, with monthly dummies from the start of the pandemic in March 2020 until the end of October 2020. A treatment effect, defined as the start of full international travel restrictions, was also included in the two estimated equations (per capita cases and per capita fatalities).

## Results

### Trend comparisons

Figure 1a and 1b, respectively, provide total cases and new cases of COVID-19 in all five countries as percentage of the respective national populations. In terms of per capita total cases, Sweden has the highest confirmed case rate that is approaching 1% of the population. This rate is about ten larger than the rate in New Zealand, which has the lowest per capita cases. Sweden’s number of cases per capita is twice as large as the next highest country, Denmark.

**Figure 1:**
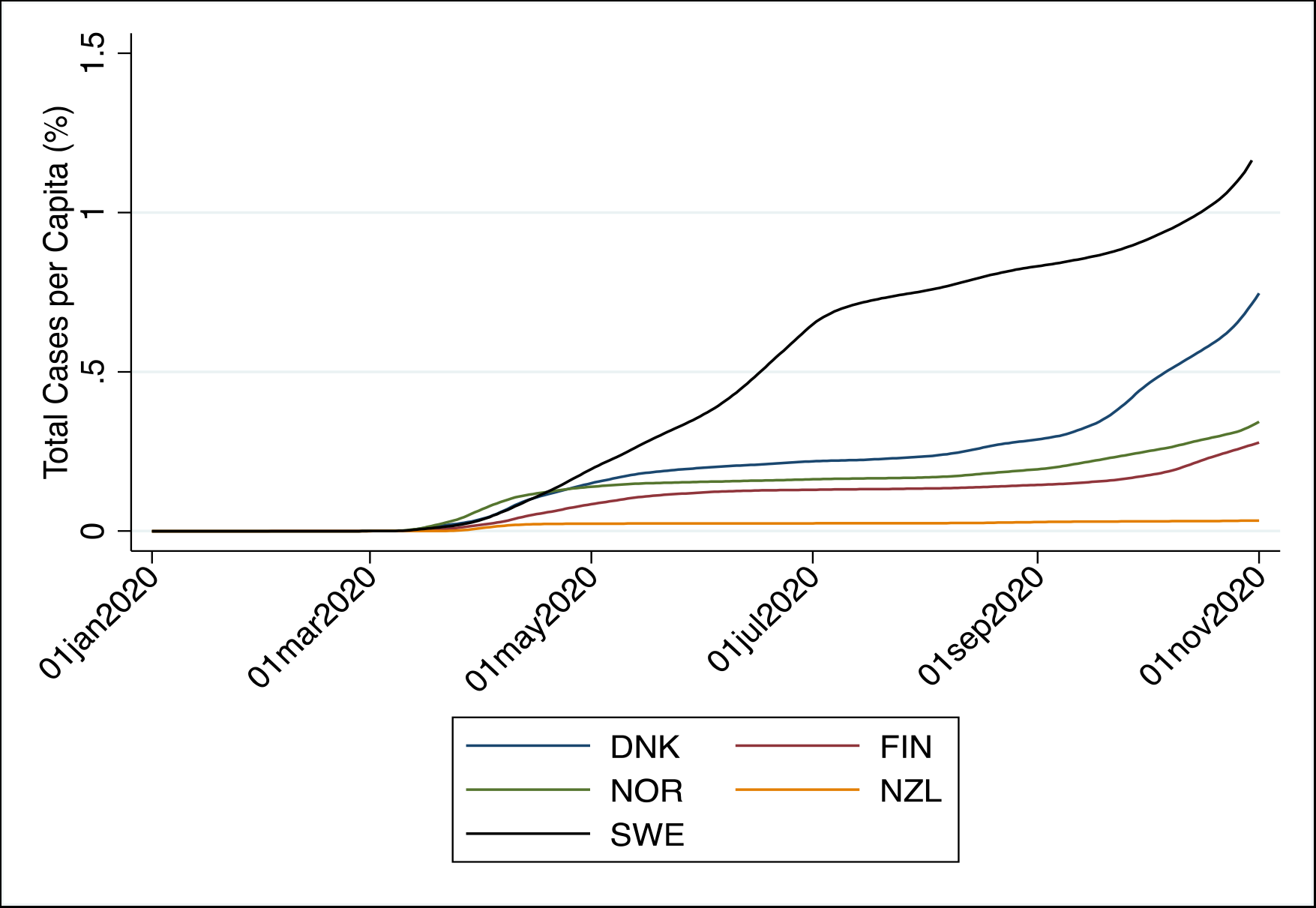
Total Cases per Capita (%): Denmark, Finland, Norway, New Zealand and Sweden (seven-day average)

**Figure 1b:**
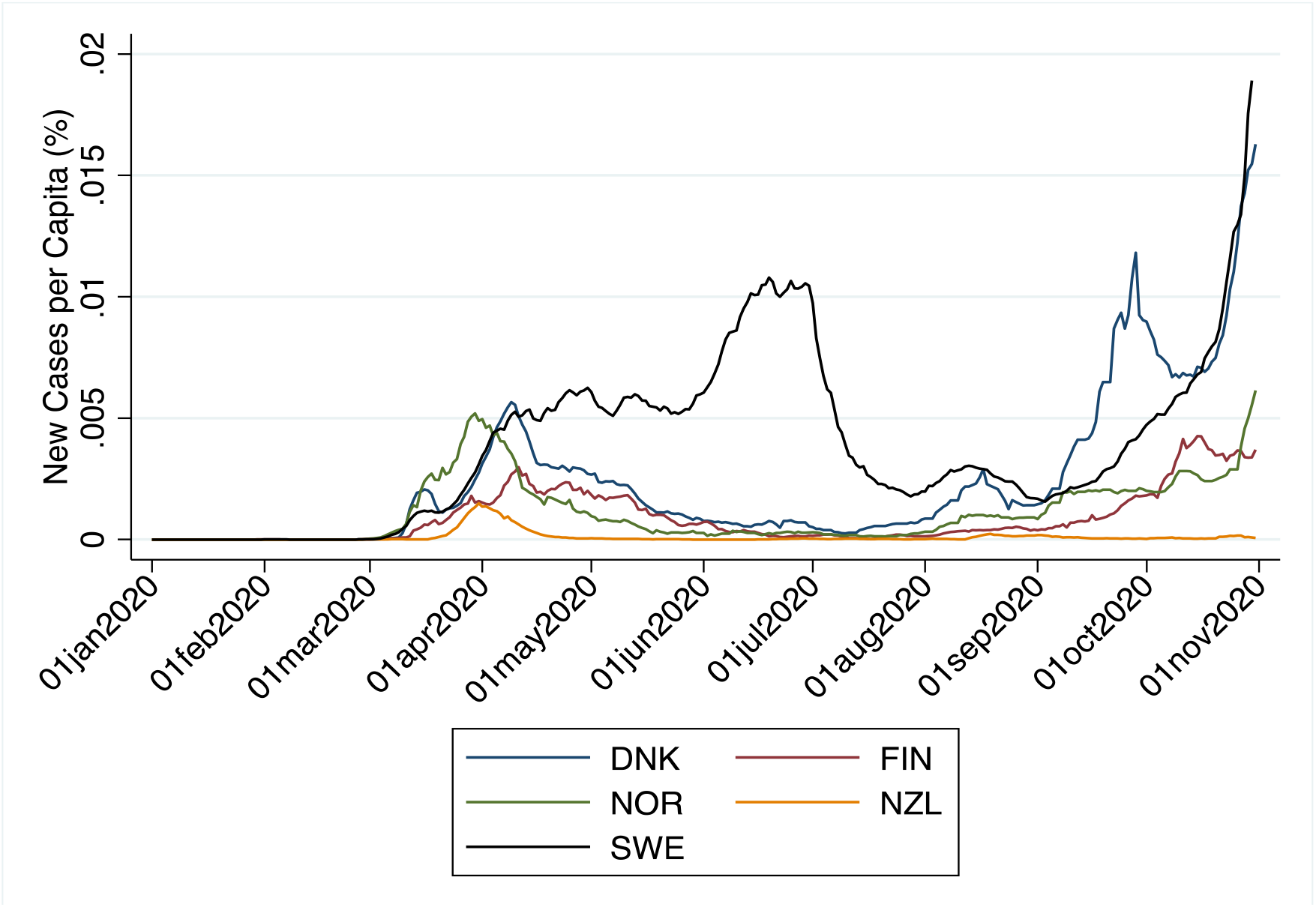
New Cases per Capita (%): Denmark, Finland, Norway, New Zealand and Sweden (seven-day average)

In terms of new cases per capita, Sweden had a much larger per capita rate of cases from April to July 2020 that exceeded 0.01% until it started to decline and become comparable with its Nordic neighbours in August 2020 at around 0.0025%. From early September 2020 to the end of October 2020, Denmark has had the highest new cases per capita of all countries since the start of the pandemic, at 0.012%. Per capita new cases in Sweden also increased in September and October 2020 and were a similar level to Denmark at the end of October 2020.

Figure 2a and 2b, respectively, provide total fatalities and additional fatalities (seven-day average) per capita as a percentage of the respective national populations. Sweden’s total fatalities per capita are some 0.06% and are approximately six times higher than the next highest per capita fatality rate in Denmark. Sweden’s per capita additional fatalities peaked in mid-April 2020 at 0.001% at about 5 times of the next highest country, Denmark. For all countries, additional per capita fatalities fell from their April peaks to August 2020, with a very slight upward trend exhibited in Denmark in September/October 2020.

**Figure 2:**
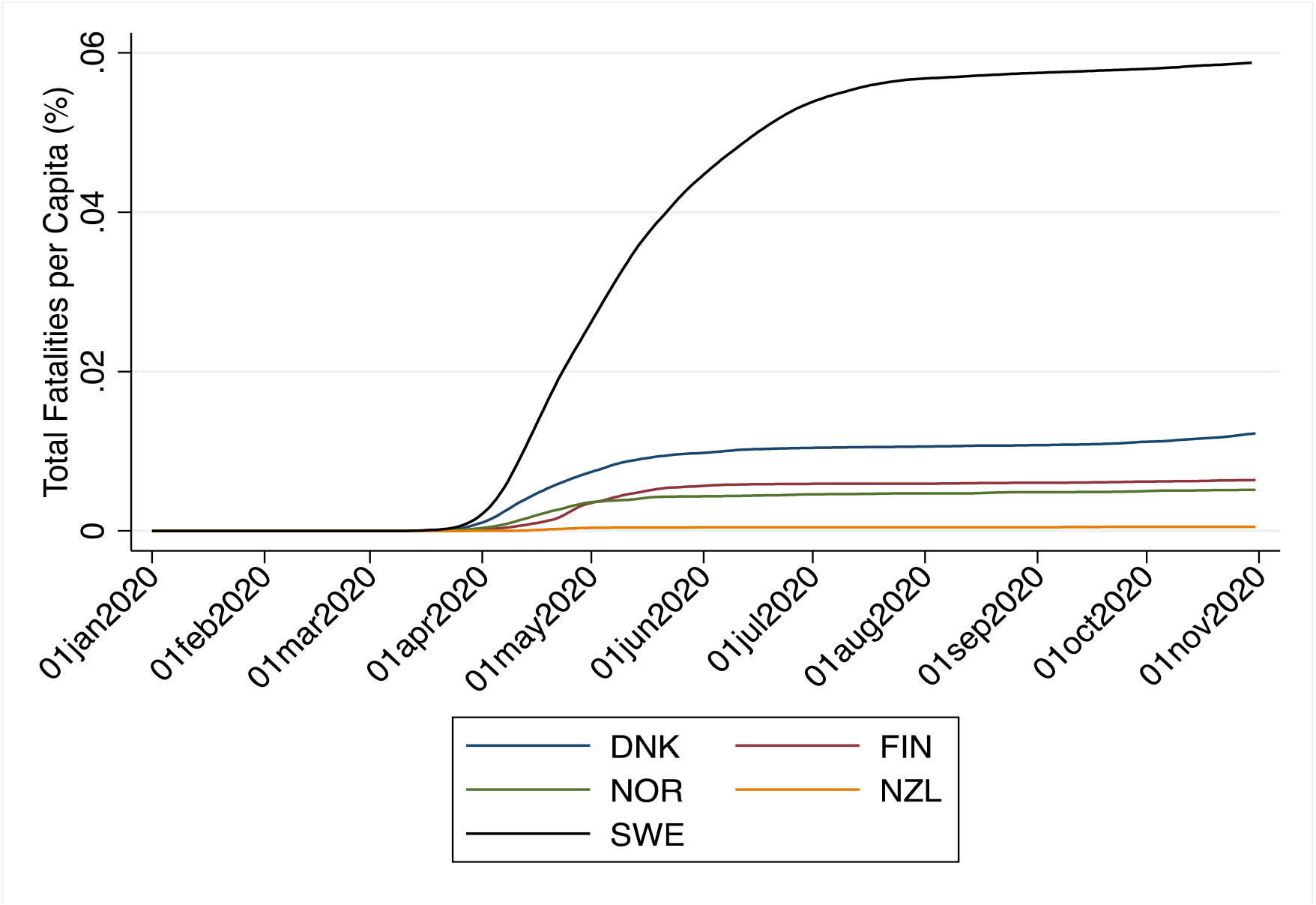
Total Fatalities per Capita (%): Denmark, Finland, Norway, New Zealand and Sweden (seven-day average)

**Figure 2b:**
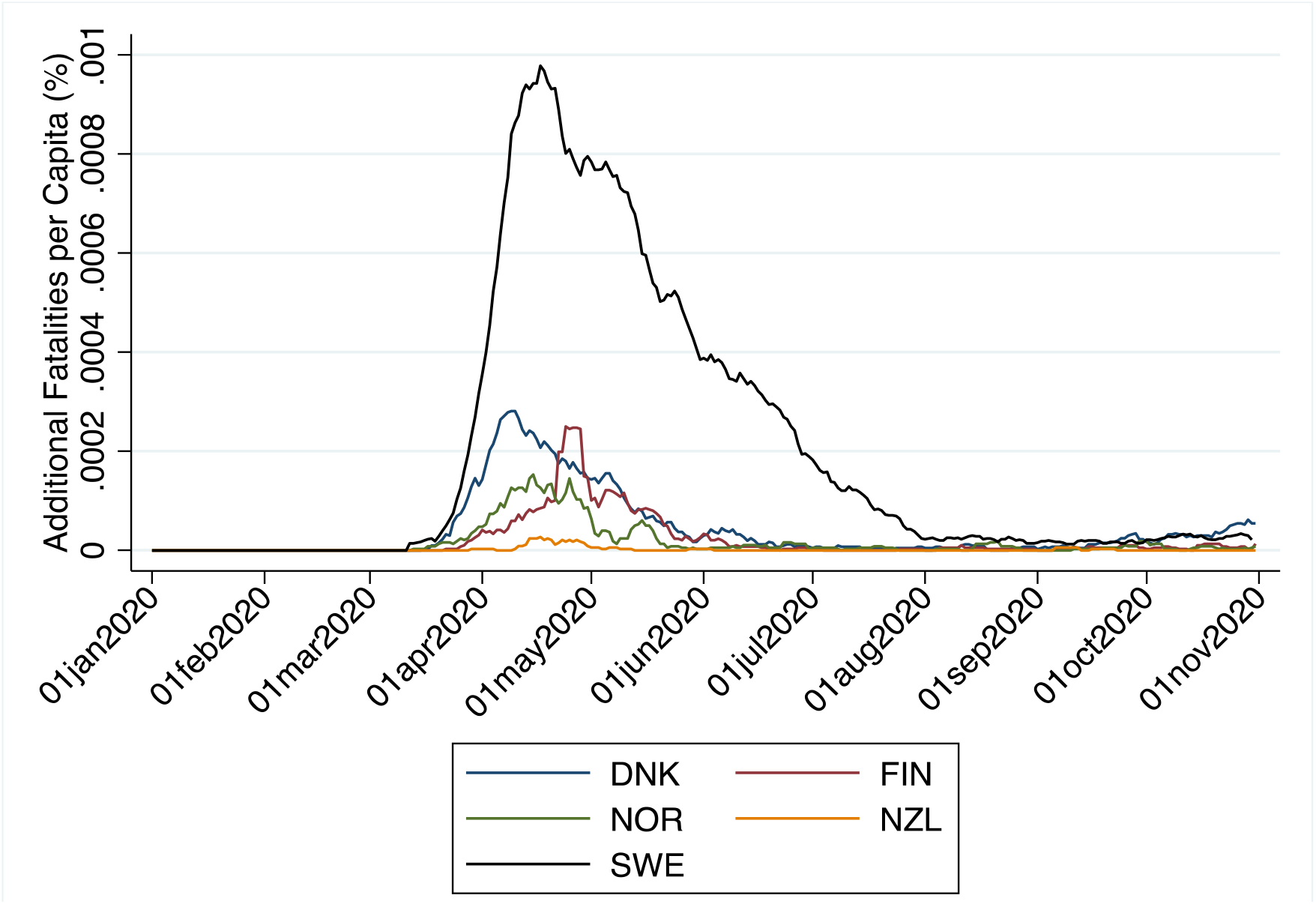
Additional Fatalities per Capita (%): Denmark, Finland, Norway, New Zealand and Sweden (seven-day average)

A key public health response to the pandemic is testing and contact tracing of confirmed cases. Figure 3a and 3b provide, respectively, cumulative tests per confirmed cases and additional tests (seven-day average) per additional confirmed cases. Sweden has had much lower testing (in total and additional), at about one hundredth of the rate of any of its Nordic neighbours and New Zealand. Since July 2020 the testing rate (total and additional) increased substantially in Sweden but it remains a small fraction of the level in the four other countries.

**Figure 3:**
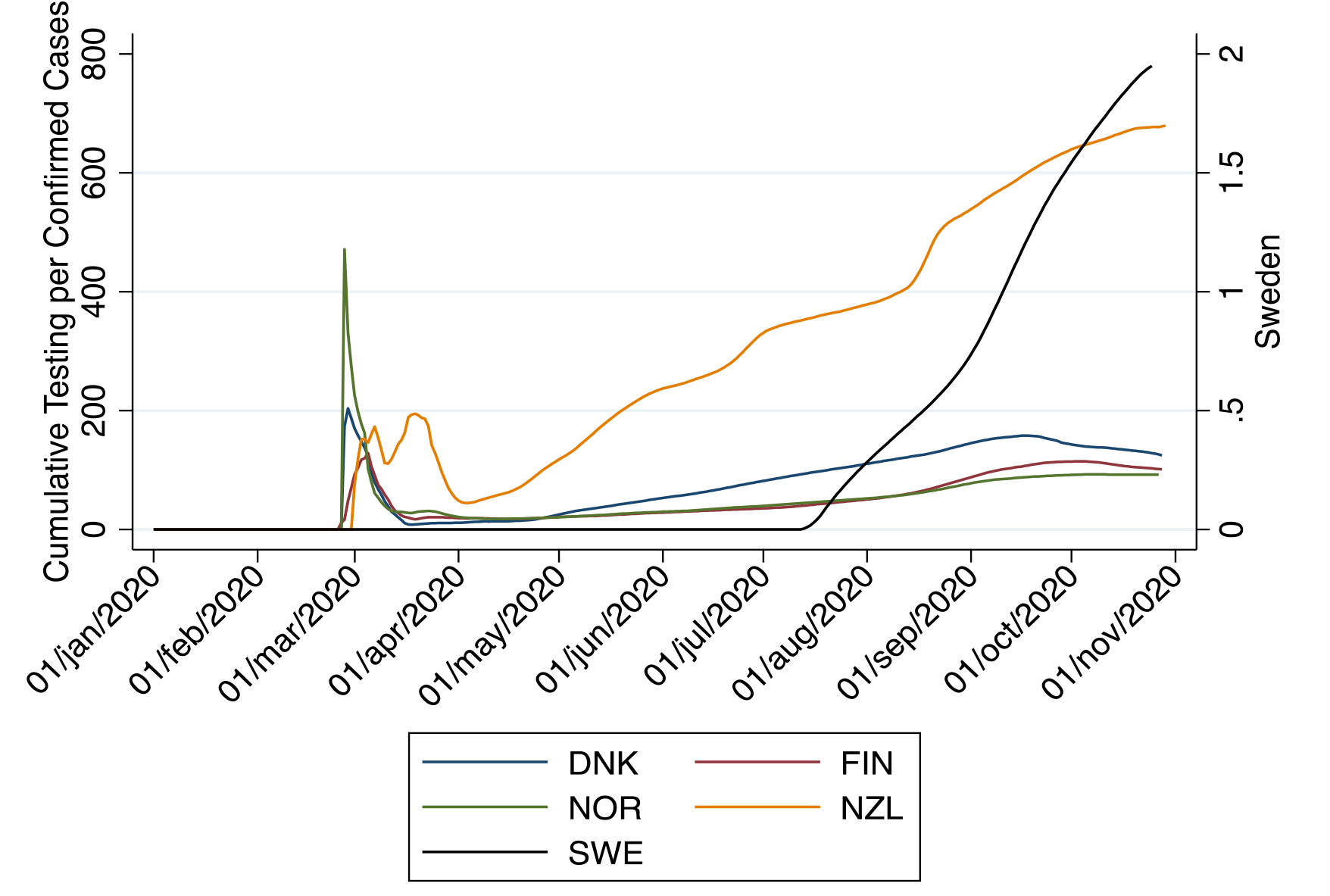
Cumulative Testing per Confirmed Cases: Denmark, Finland, Norway, New Zealand and Sweden (seven-day average)

**Figure 3b:**
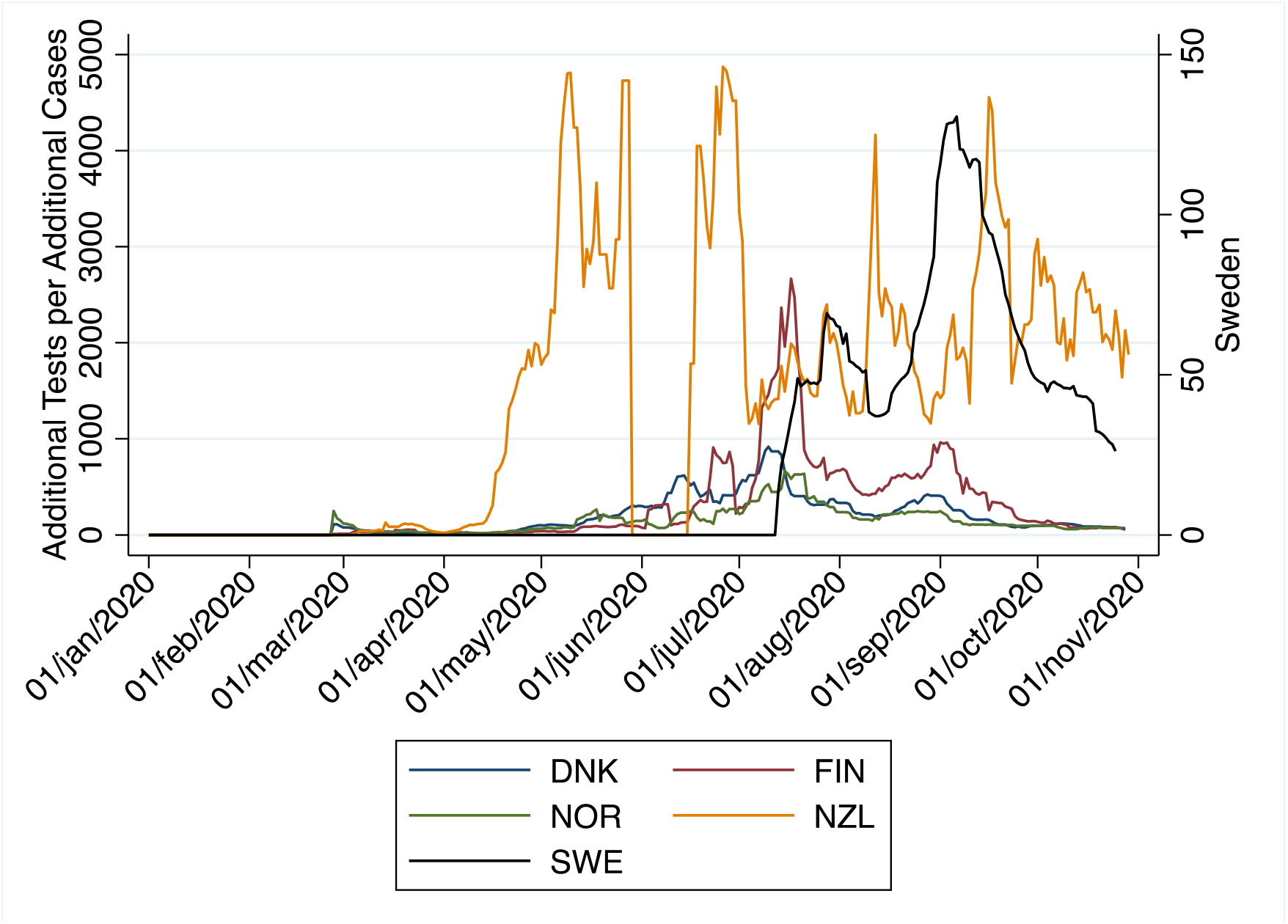
Additional Tests per Additional Confirmed Cases: Denmark, Finland, Norway, New Zealand and Sweden (seven-day average)

Figure 4 provides a level (0 to 100) of the stringency of government-mandated SD measures using an aggregated of the sub-indices C1-C8, E1 and H1-H3 from the University of Oxford Coronavirus Government Response Tracker (for details see https://www.bsg.ox.ac.uk/research/research-projects/coronavirus-government-response-tracker), with 100 being the highest level of stringency. New Zealand had the highest level of stringency in its first outbreak in March and May at levels approaching 100. By contrast, Sweden had the lowest stringency level over the March-May period of between 40 and 50, Norway had stringency levels as high as 80, and Denmark and Finland had levels, respectively, of around 70 and 60.

**Figure 4:**
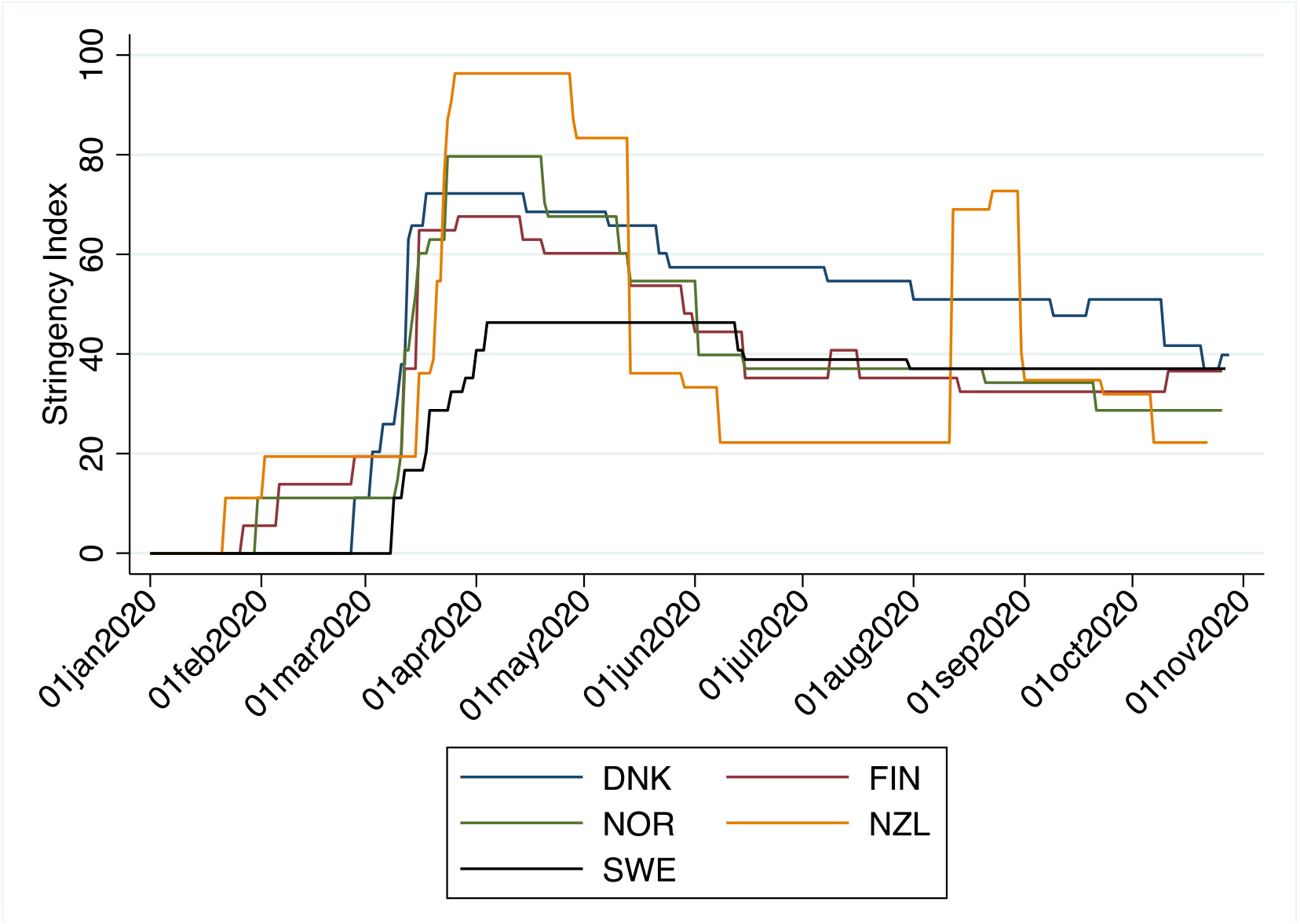
Stringency Index: Denmark, Finland, Norway, New Zealand and Sweden (seven-day average)

Stringency levels declined for all countries from May 2020 onwards and fell the most for New Zealand that had the lowest stringency levels of all at around 20 by June 2020 following its elimination of community transmission of COVID-19. New Zealand’s stringency levels rose quickly and approached 80 in August with a second outbreak but fell again in September after the outbreak was successfully contained and community transmission eliminated.

Sweden’s stringency level has changed the least during the pandemic and has remained at around the 40 level since April 2020. Its stringency levels over the June-September 2020 period were similar to Finland and Norway. With the exception of the spike in stringency levels in New Zealand in August 2020, following its second outbreak, Denmark maintained the highest stringency level from May 2020 at around 50 but it decreased to Swedish levels by October 2020.

Figure 5 provides the percentage quarterly change in GDP, a measure of economic performance, from the fourth quarter of 2019 to the end of the second quarter of 2020 and the corresponding total quarterly number of COVID-19 fatalities (given by the number in the square bracket for each quarterly bar) for Denmark, Finland, Norway, New Zealand, and Sweden. With the exception of Sweden that has about 10 million residents, all other countries have populations between five and six million people.

**Figure 5:**
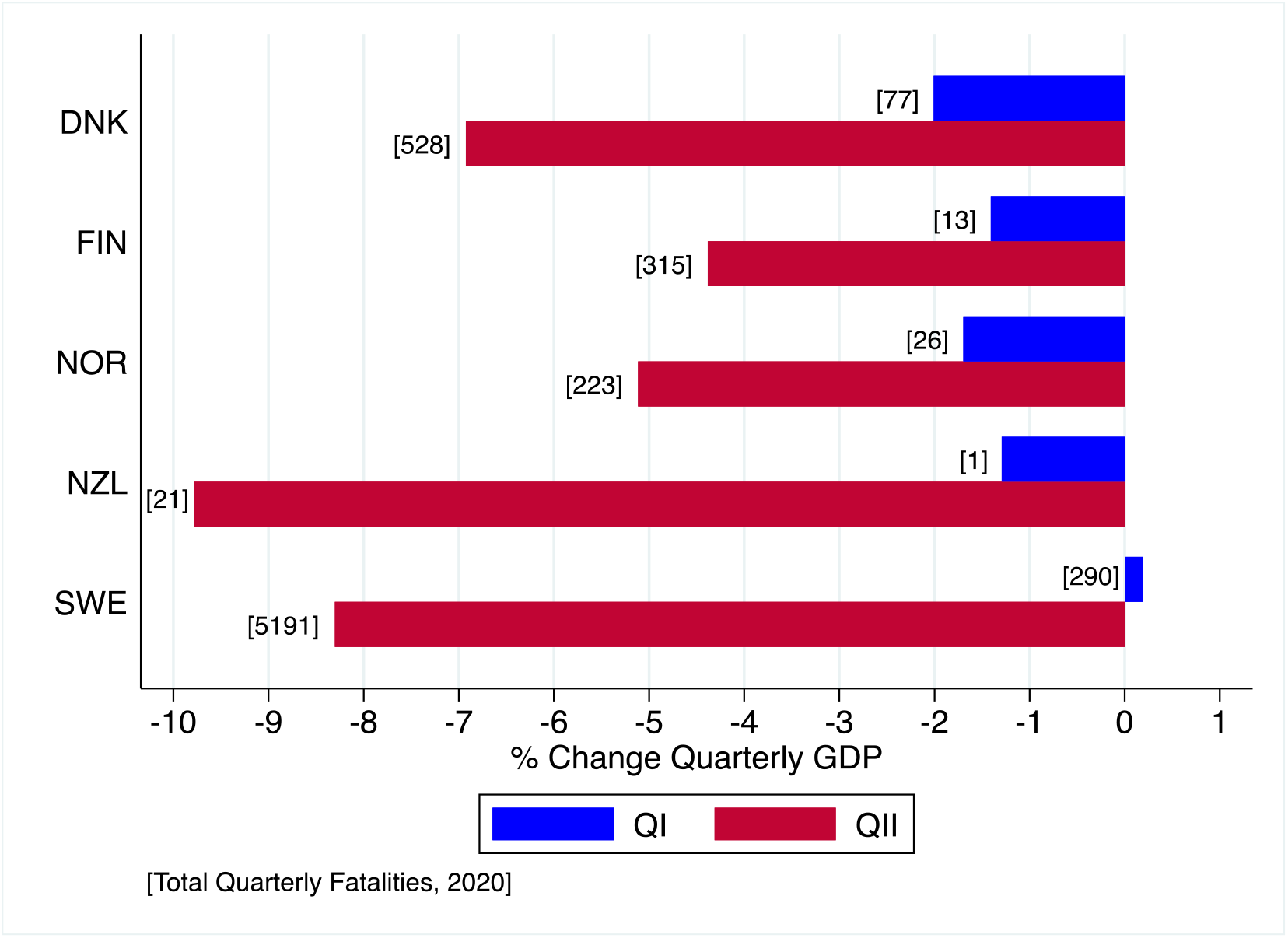
Percentage Change in Quarterly GDP with Total Quarterly Cumulative Fatalities: Denmark, Finland, Norway, New Zealand and Sweden

In each quarter, New Zealand had by far the lowest number of cumulative COVID-19 fatalities and total fatalities to the end of October of less than 30. Sweden had the highest quarterly fatalities with total fatalities (cumulative over both quarters) approaching 5,500 and a cumulative total, as of the end of October 2020, of almost 6,000. Sweden’s Nordic neighbours reported much lower quarterly fatalities and all had total fatalities at the end of October 2020, not exceeding 750. The change in GDP between the first and second quarters is negative for all countries and varies from minus 4% and 5% for Finland and Norway, about minus 7% for Denmark, about minus 8% for Sweden, and almost minus 10% for New Zealand.

### Difference-in-Difference results

The complete DiD results for per capita confirmed cases and per capita fatalities are available from the authors on request. Summary results of the estimated coefficients (with monthly fixed effects) and their *p*-values for the two DiD regressions are provided in Equations (1) and (2):

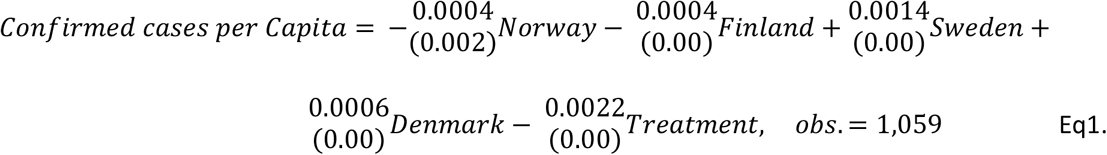

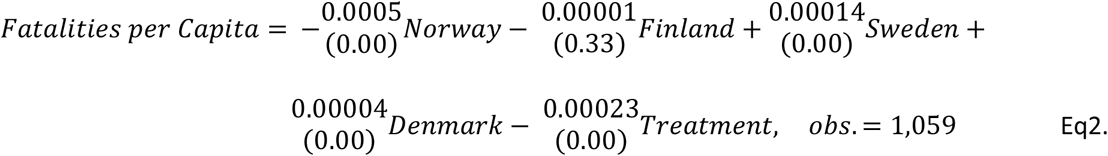

A statistical test for differences across the four Nordic countries over the period March to September 2020 indicates that both Denmark and Sweden had a significantly positive country effect on confirmed per capita cases and per capita fatalities. Thus, relative to the reference country (Norway), both Denmark and Sweden had statistically significant higher levels of per capita confirmed cases and per capita fatalities. The magnitude of this difference, however, was much higher (more than ten times larger) in Sweden than in Denmark.

The treatment effect of full international travel restrictions is statistically significant from zero at the standard level of significance. In sum, the results imply: (1) that border closures, at least in Nordic countries, in the first half of 2020 *reduced* both confirmed cases and fatalities associated with COVID-19 and (2) Sweden, by far, had the largest country effect on *increased* per capita cases and per capita fatalities of the four Nordic countries.

## Discussion

### Main findings

While variations exist among the four countries, including in terms of trust in government [15], the selection of neighbouring Nordic countries ensured, as much as possible, like-with-like national comparisons and reduced the effects of key cross-country differences such as quality of health care, welfare payments, per capita income, capacity of government, education, demography and geography. Based on analysis of trends from 1 January to 31 October 2020, we find that Sweden’s public health outcomes, in terms of per capita cases and fatalities from COVID-19, are much worse than in its Nordic neighbours.

Based our trend analysis of the level of stringency of government-mandated SD until May 2020, we found that the different public health response adopted by Sweden, with a focus on voluntary SD rather than government-mandated SD, is a plausible explanation for cross-country differences in public health outcomes (per capita cases and per capita fatalities). Another plausible factor for the difference among the selected countries is that Sweden has had a much lower rate of testing for COVID-19 compared to its peers. A lower testing rate reduces the efficacy of contact tracing that, in turn, contributes to higher number of infections [16] for any given level of SD (mandated or voluntary).

A claimed benefit of voluntary SD, as practised in Sweden, was it would provide for a higher level of economic activity compared to more stringent and government-mandated SD. Cross-country economic comparisons are problematic in that factors, other than public health outcomes, may have contributed to a better or worse performance. Nevertheless, we do not find evidence for the proposition that the reliance on voluntary SD by Sweden has resulted in superior economic performance, as measured by change quarterly GDP growth, relative to its peers.

### Limitations

The principal limitation of our study is that the pandemic will continue for many more months, possibly even years, even with the widespread use of vaccines [17]. Thus, our findings of a worse public health performance, and no better economic performance, in Sweden relative to its Nordic neighbours only applies to the end of October 2020. A second limitation is that because we have adopted a ‘like-with-like comparison’ to control for cross-country differences we have only five observations at a country level. In turn, this limits our ability to undertake statistical tests for differences in means among the selected countries. A third limitation is that the data on confirmed cases of COVID-19 are likely to be less than actual cases [18] and this is likely to more pronounced in countries that have had a low testing rate, such as Sweden. A downward bias may also apply, at least in some countries, in relation to COVID-19 fatalities [12]. Another data limitation is that our stringency measure of SD only accounts for government-related measures and does not account for the voluntary behaviours of individuals themselves to avoid becoming infected, or infecting others, with the SARS-CoV-2 virus.

### Implications

Notwithstanding the limitations of our study, an important public health implication of our findings is that early imposition of full international travel restrictions combined, with high levels of government-mandated stringency levels of SD, appear to have reduced the per capita cases and per capita fatalities associated with COVID-19 in Nordic countries in 2020. An economic implication of our findings is that adopting less stringent government-mandated SD is *not* associated with higher quarterly economic growth, at least in the four selected Nordic countries.

## Data Availability

All data used are publicly available and available on request.

https://econ.ucalgary.ca/profiles/daniel-gordon

## Acknowledgements

None.

